# Early life attachment in term and preterm infants

**DOI:** 10.1101/2022.04.29.22274482

**Authors:** Lorena Jiménez-Sánchez, Lorna Ginnell, Sinéad O’Carroll, Victoria Ledsham, Amy Corrigan, Yu Wei Chua, David Q. Stoye, Gemma Sullivan, Jill Hall, Ann M. Clemens, James P. Boardman, Sue Fletcher-Watson

**Affiliations:** Translational Neuroscience PhD programme, Centre for Clinical Brain Sciences, The University of Edinburgh, Edinburgh, UK; Salvesen Mindroom Research Centre, The University of Edinburgh, Edinburgh, UK; MRC Centre for Reproductive Health, The Queen’s Medical Research Institute, The University of Edinburgh, Edinburgh, UK; Centre for Discovery Brain Sciences, University of Edinburgh, Edinburgh, UK

**Keywords:** Attachment, prematurity, infancy, socioeconomic deprivation

## Abstract

1

**Background:** Preterm birth is associated with atypical cognitive and socioemotional outcomes in childhood. Secure infant attachment protects against adverse outcomes, but could be modified by alterations in the early caregiving environment inherent to essential neonatal intensive care or co-morbidities of preterm birth. We aimed to test the hypothesis that preterm birth is associated with differences in infant attachment, and to investigate clinical, neurodevelopmental and socioeconomic variables that are associated with infant attachment.

**Methods:** 82 preterm and 75 term infants with mean (range) gestational age at birth 29.5 (22.1 – 32.9) and 39.6 (36.4 – 42.1) weeks, respectively, completed the Still-Face Paradigm (SFP) at nine months of corrected age. Attachment dimensions and categories were obtained from infant responses to the SFP during the reunion episode using a published coding scheme, and an alternative principal component (PC) and clustering strategy. Neurodevelopment was assessed using the Vineland Adaptive Behavior Scales, and socioeconomic status was operationalized as neighborhood deprivation.

**Results:** Preterm and term infants significantly differed in fretfulness, attentional PC scores and in their distribution between attachment clusters (p-values ≤ 0.3); with preterm infants exhibiting less fretful and more neutral responses to the SFP. Preterm and term infants did not significantly differ in distress, attentiveness to caregivers, emotional PC scores, or in their distribution between attachment styles (p-values ≥ .13). In the whole sample, fretfulness correlated with socioeconomic deprivation (r_s_ = −0.18, p-value = .02).

**Conclusions:** Data reveal subtle attachment differences between preterm and term infants at nine months of age, which may not always be captured by traditional approaches for categorizing attachment. Findings suggests that caregiver-infant attachment relationships may not be fully resilient to the effects of prematurity on the developing infant, but this depends on how attachment is measured. Our results highlight putative links between socioeconomic deprivation and infant attachment that warrant further study.

## 4 Introduction

Preterm birth (birth before 37 weeks of gestation) is associated with atypical neurodevelopmental outcomes in childhood, including attention problems, impaired language development and delayed socioemotional competence (Ene et al., 2019, Dean et al., 2021a, Dean et al., 2021b, Barre et al., 2011). Secure infant attachment can protect against some of these adverse outcomes (Thompson, 2008); however, it could be adversely modified by unavoidable alteration in the caregiving environment inherent to neonatal intensive care required by preterm infants. These alterations include an inevitable degree of separation from parental caregivers, infant stress, and co-exposures of preterm birth that affect the developing brain (Korja et al., 2012, Boardman and Counsell, 2020). Infant attachment could also be impacted by atypical cognitive capacities linked to preterm birth, for example, differences in attention maturation (Ginnell et al., 2021) may influence how long preterm infants engage in social interactions (Burstein et al., 2021). A better understanding of the relationship between prematurity and attachment is essential to support early relationships between preterm infants and their caregivers, which in turn, are important to foster infants’ socioemotional resilience and parental wellbeing.

Early interactions between infants and their caregivers build infants’ attachment relationships (Ainsworth et al., 1972, Emde, 1980) and shape infants’ attachment style (Bowlby, 1982). Securely attached infants use caregivers as a secure base to explore from and return to. Under a threat, insecurely attached infants either avoid their caregiver (avoidant) or fail to respond independently (resistant). Secure, avoidant and resistant attachment styles are adaptative organized strategies, but some infants show contradictory behaviors i.e. attachment disorganization (Main and Solomon, 1990). Secure attachment has been shown to contribute positively to infants’ socioemotional development (Thompson, 2008), increased peer competence (Groh et al., 2014) and mental health in later life (Spruit et al., 2020). For preterm infants specifically, secure attachment associated with a better neurological development at 14 months (Brisch et al., 2005) and higher cognitive scores at 24 months of corrected age (López-Maestro et al., 2017). Attachment has also been linked to demography: secure preterm infants were less likely to come from lower socioeconomic backgrounds (Wille, 1991).

Research has examined whether infant attachment is disrupted by preterm birth, but findings are inconsistent. Some studies found no difference in attachment styles between preterm (gestational age at birth (GA) < 37 weeks) and term infants (Rode et al., 1981, Frodi and Thompson, 1985, Goldberg et al., 1986, Plunkett et al., 1986, Easterbrooks, 1989, Butcher et al., 1993, Brisch et al., 2005). Other studies reported that preterm infants of GA 32 – 36 weeks were more frequently assigned an avoidant attachment style (Fuertes et al., 2022), preterm infants of GA < 32 weeks were more commonly assigned a disorganized attachment style (Wolke et al., 2014), and preterm infants of GA < 26 weeks were more frequently assigned a resistant attachment style (López-Maestro et al., 2017, Sierra-García et al., 2018). All the aforementioned studies followed similar methodologies and assessed attachment from 12 months of age, but studies demonstrating differences in attachment were more recent, and included larger cohorts of preterm infants with a lower mean GA. Neonatal Intensive Care Unit (NICU) practices and outcomes have substantially evolved since the 1990s, resulting in higher survival rates of extremely preterm infants and better developmental outcomes (Bell et al., 2022). Therefore, there is a need to explore the relationship between prematurity and attachment in contemporary cohorts of recipients of modern intensive care. Contradictory findings may also be explained by the caregiver’s gradual adaptations to the characteristics of the preterm infant during the first year of life, establishing a more harmonious interactional style suited to the infant (Frodi and Thompson, 1985). Thus, if preterm infants do present differences in attachment, these could be more apparent early in infancy.

Early attachment has traditionally been studied using the Strange Situation Procedure (Ainsworth et al., 1972), where toddlers are observed playing while caregivers and strangers enter and leave one room. The Strange Situation assumes infants’ motor competence and fear of strangers, characteristics that may not be completely developed in those younger than 12 months (Schaffer, 1966), especially in clinical populations. The Still-Face Paradigm (SFP) offers an opportunity to study younger populations, while infants progress in terms of locomotor ability and intentionality (Zeanah et al., 1989) and become less indiscriminately friendly and more wary of strangers (Schaffer, 1966). From seven-to-eight months of age, these developments go hand-in-hand with the formation of attachment to a primary caregiver (Abbott, 2016). Current approaches for categorizing attachment styles from the SFP use predefined thresholds based on raw scores that capture infants’ emotions and attentiveness to the caregiver (Williams and Turner, 2020, Abbott, 2016). These approaches may not be appropriate for preterm infants who may have atypical attention profiles. Data-driven approaches can partially address this limitation by categorizing attachment based on the behavioral heterogeneity of the population. Moreover, infant attachment patterns may be more continuously rather than categorically distributed, so broad approaches exploring both attachment dimensions and categories are recommended (Fraley and Spieker, 2003).

Our primary aim was to test the hypothesis that preterm birth is associated with differences in infant attachment using two methods to analyze infant behaviors coded from the SFP: an existing SFP categorization system and a sample-specific data-driven approach. A secondary aim was to investigate clinical, neurodevelopmental and socioeconomic variables that are associated with infant attachment in the whole sample.

## 5 Methods

### 5.1 Participants

Participants were part of a longitudinal cohort study designed to investigate the effect of preterm birth on brain development and neurocognitive outcomes (Boardman et al., 2020). Preterm (GA < 33 weeks) and term infants were born between October 2016 to March 2021 at the Royal Infirmary of Edinburgh, UK. Preterm infants transferred to the hospital *ex utero* for intensive care, and infants with congenital anomalies were excluded. Informed written parental consent was obtained. Ethical approval was obtained from the National Research Ethics Service (16/SS/0154), South East Scotland Research Ethics Committee, and NHS Lothian Research and Development (2016/0255).

Caregivers were invited to attend follow-up appointments with their infants at nine months of age. Corrected age was used for the preterm group. Participants attended the Child Development lab, part of the Salvesen Mindroom Research Centre and the University of Edinburgh, at the Royal Edinburgh Hospital. The visit comprised an assessment battery including questionnaires and behavioral procedures (for protocol, see Boardman et al., 2020).

Demographic data were collected through questionnaires and review of medical records. Collected information included sex, GA, birth weight (grams), total number of days spent in the NICU, age at testing (months), and the Scottish Index of Multiple Deprivation 2016 (SIMD) rank, generated from postcode information collected via parental questionnaire. SIMD rank (Scottish Government, 2016) is a multidimensional score generated by the Scottish government ranking localities’ deprivation according to local income, employment, health, education, geographic access to services, crime and housing.

### 5.2 Measures and materials

#### 5.2.1 The Still-Face paradigm

After video consent was obtained, infants were secured in a high chair. Caregivers seated facing the infant approximately 50 cm away. Two Panasonic HC-W580 video cameras were set up behind the caregiver and the infant to record the infant and caregiver’s faces and hands. All researchers seated out of view, and verbally cued caregivers. The procedure included five two-minute episodes (Haley and Stansbury, 2003); modified from the original protocol by (Tronick et al., 1978): baseline, still-face, reunion, still-face, and reunion. During the baseline episode, caregivers were instructed to interact naturally with the infant, without using toys. During still-face episodes, caregivers were instructed to express a neutral facial expression, remaining still and looking slightly above the infant’s head, avoiding eye contact and interaction with their infant. During reunion episodes, caregivers were instructed to interact normally with the infant again, without toys. Caregivers were always given the option to terminate the paradigm if their infant exhibited severe distress.

#### 5.2.2 Data coding

Williams and Turner’s coding scheme (Williams and Turner, 2020) was selected to study attachment from the SFP at nine months of age. Williams and Turner’s coding scheme includes the Happy-Distressed (HD), Not fretful-Fretful (NFF) and Attentive-Avoidant (AA) Global Rating Scales (GRS) from Murray et al. (1996) to analyze infant behaviors during the reunion episode (Abbott, 2016, Williams and Turner, 2020). The coding scheme was applied as per the coding manual. To study attachment dimensions, raw scores of each scale were calculated as the percentage of time the infants engaged in each scale’s set of behaviors during the reunion episode, offering continuous information of the HD, AA and NFF scales. To study attachment categories, attachment styles were obtained according to the published algorithm. In this study, only the first reunion episode of the five-episode SFP was coded because this provided a better equivalent to previous studies, where infants only experienced one stress episode (the still-face episode) before the reunion (Abbott, 2016, Williams and Turner, 2020).

Video coding was conducted using EUDICO Linguistic Annotator software (Wittenburg et al., 2006).

Participants were excluded based on SFP procedural violations (cases where others than the caregiver-infant dyad initiated a violation), and coded reunion time (cases where the duration of the codable time during the reunion episode < 30 seconds, see Appendix S1 for further details).

#### 5.2.3 Assessment of development

Infants’ behavioral development was assessed by the Vineland Adaptive Behavior Scales Third Edition Comprehensive Interview Form Report (Sparrow et al., 2016, Dean et al., 2021b). Caregivers were interviewed by trained examiners to complete the communication, social, daily living skills and motor skills domains for all infants during the follow-up appointment at nine months of age. For each subdomain, the sum of raw scores and v-scale scores were extracted (Figure S1). Raw scores represent the total number of items i.e. adaptive behaviors infants present at the time of assessment, while v-scale scores are calculated by scaling raw scores (mean = 15, standard deviation = 3) according to the infant’s corrected age at the time of assessment. Sum of raw scores per subdomain were used for subsequent analyses because 1) raw scores have been recommended when ability is of interest (Farmer et al., 2020), 2) raw scores captured more behavioral heterogeneity than standard scores (Figure S1), and 3) we did not have specific hypotheses about age.

### 5.3 Data analyses

Data analyses and plots were generated using R (version 4.2.1; R Core Team, 2020). R packages used in this study are listed in Appendix S2.

#### 5.3.1 Principal component and clustering analyses

A data-driven approach was carried out to generate sample-specific data-driven attachment dimensions and categories for two reasons. First, Williams and Turner’s algorithm imposes numeric cut-offs to assign attachment styles, but behavioral differences may be subtle around those numeric cut-offs. Second, it has been debated whether attachment styles are naturally occurring groups across different populations (Fraley and Spieker, 2003). Cluster analysis can identify new behavioral groups, maximizing similarities within each group and minimizing similarities between groups (Henry et al., 2005).

A principal component (PC) analysis was performed to generate attachment dimensions for clustering. The selected variables included the total raw scores of Williams and Turner’s coding scheme (Williams and Turner, 2020). Since disorganized attachment goes along with an increase of distress across the reunion episode, HD raw scores for the first and second half of the reunion episode were also included. Thus, PC analysis input variables were total HD, first-half HD, second-half HD, total NFF and total AA raw scores.

Agglomerative hierarchical clustering was carried out on all principal components (Coppola et al., 2016). To create more compact and even sized clusters (Szmrecsanyi, 2012), the Ward’s criterion was used as linkage method and the minimized squared Euclidean distance as the distancing metric. The number of clusters with the highest relative loss of inertia was chosen. This approach involved no a priori assumption about the number of clusters, providing the maximum flexibility in determining the appropriate number of groups. Data-driven approaches can sometimes provide cluster solutions despite weak cluster separation, so it is important to determine whether cluster solutions are stable and should be retained. Cluster stability was assessed by bootstrapping, comparable to Matsushita and colleagues’ approach (2022): in-silico replicated datasets were generated from the original dataset and hierarchical clustering was performed in all replicates (B = 500 iterations; see Appendix S3 and Figure S2 for further details). Alternative clusters were compared to original clusters by computing the Rand index values for each pair of clusters on each iteration, and the matrix of the average Rand index values across iterations for each pair of clusters was reported.

#### 5.3.2 Statistical analyses

Distributions of attachment categories in the term and preterm groups were compared using the chi-square test. For chi-square tests, effect sizes were calculated with Cramér’s V (Cramér, 2016). To further investigate the relationship between prematurity and individual attachment categories, multinomial logistic regression was used with attachment categories (attachment styles or attachment clusters) as outcomes and prematurity (preterm vs term-born) as predictor. To analyze attachment dimensions, Shapiro-Wilk tests and F-tests were used to assess data normality and homogeneity of variances, respectively. Attachment dimensions between preterm and term infants were compared using t-tests or Mann-Whitney-Wilcoxon tests for normally or non-normally distributed data, respectively. For t-tests, effect sizes were calculated with Cohen’s d. For Mann-Whitney-Wilcoxon tests, effect sizes were calculated with r (rank-biserial correlation; Rosenthal et al., 1994). Correlations between attachment dimensions and demographic factors or behavioral development were investigated using Pearson or Spearman’s rank correlation coefficient for normally or non-normally distributed data, respectively. Since correlations were exploratory, Pearson or Spearman’s rank p-values were not corrected (Armstrong, 2014).

A post-hoc power analysis for PC and clustering attachment data in the study sample size was conducted. For attachment clusters, the behavioral sample provides 90% power to detect a medium effect size (.28) at an alpha-level of .05 (χ^2^). For attachment PC scores, the behavioral sample provides 90% power to detect a medium effect size (.35) at an alpha-level of .05 (Mann-Whitney-Wilcoxon test using Shieh and colleagues’ approach, 2006).

Unless otherwise specified, results are presented as mean (standard deviation, SD) for parametric data and median (range) for non-parametric data, with significance threshold set at p-value < .05.

#### 5.3.3 Confounder analyses

Attachment outcomes that significantly differed between preterm and term infants were compared adjusting for possible confounders (i.e. variables that significantly correlated with any attachment dimension). To compare attachment dimensions between preterm and term infants adjusting for confounders, generalized linear regression was used with attachment dimensions as outcomes, prematurity (preterm vs term born) as predictor, and confounders as covariates (i.e. additive predictors). To compare attachment categories between preterm and term infants adjusting for confounders, multinomial logistic regression was used with attachment categories as outcomes, prematurity (preterm vs term born) as predictor, and confounders as covariates (i.e. additive predictors).

#### 5.3.4 Reliability analyses

LJ-S coded all videos, and LG coded 10 % of all available videos for reliability analyses (Figure 1). The main coder was blind to group (preterm vs term infants). Both researchers independently rated all of the infant behaviors on the HD, NFF, AA scales following Williams and Turner’s coding manual (2020). Two types of inter-rater reliability scores were calculated. For attachment dimensions (raw scores on the HD, NFF and AA scales), the intraclass correlation coefficient (ICC) was calculated. Following Williams and Turner’s coding scheme, two coders would need to agree on the absolute values of the HD, NFF, and AA raw scores to assign the same attachment style to a subject. Thus, a two-way random-effects agreement ICC was calculated for each scale. For attachment categories (attachment styles), the percentage of agreement (tolerance = 0) and Cohen’s Kappa were calculated.

**Figure 1.**
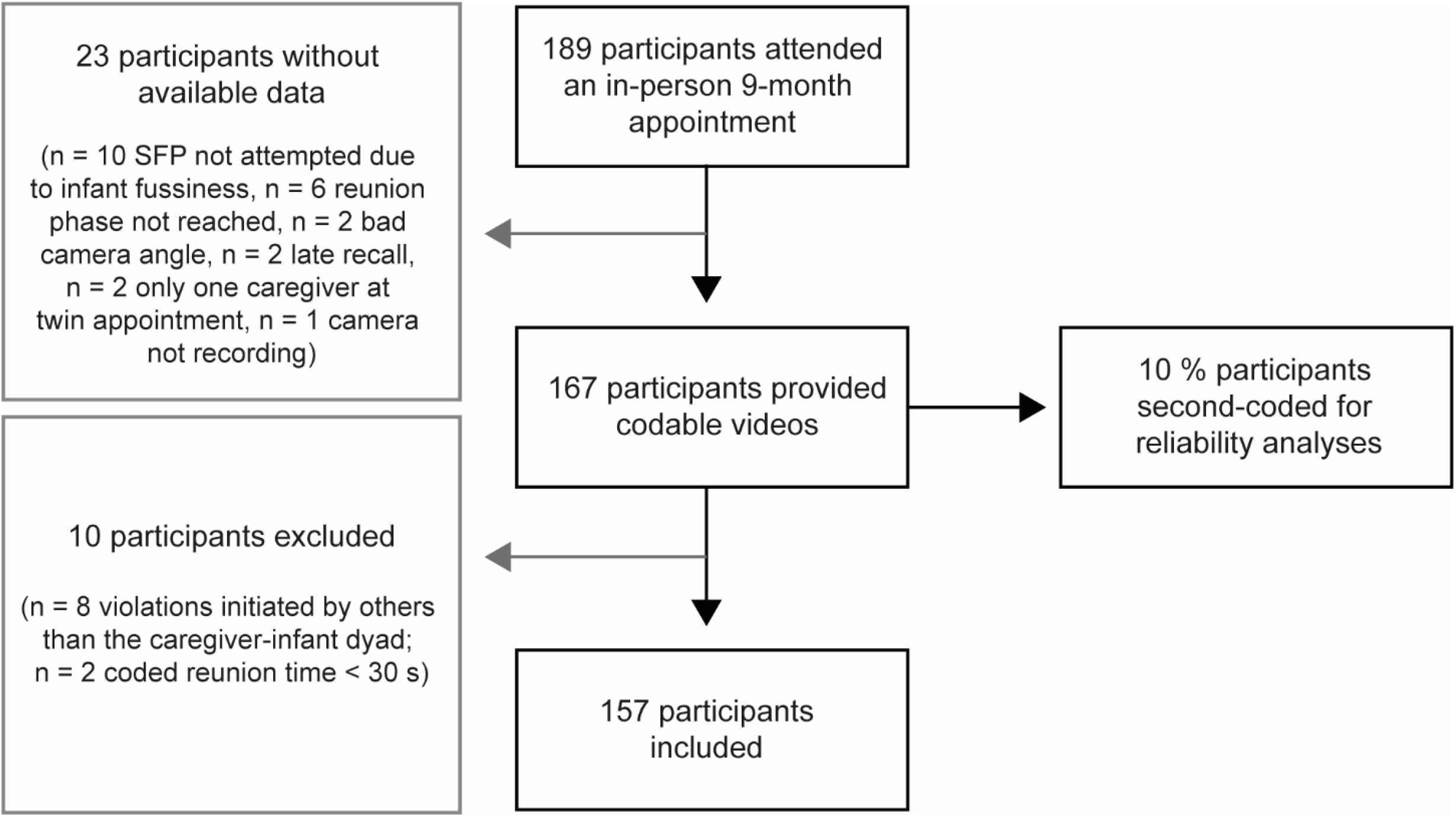
Study sample inclusion and exclusion flowchart.

ICC was .92 for the HD raw scores, .86 for the NFF raw scores and .81 for the AA raw scores. The percentage of agreement was 76.50 % and Cohen’s kappa was .65 for attachment styles.

### 5.4 Data availability

Data availability is governed in accordance with the TEBC data access and collaboration policy, which is available on request

## 6 Results

### 6.1 Baseline characteristics

One hundred and eighty nine caregiver-infant dyads completed in-person follow-up appointments at nine months, of which 167 provided codable videos and were assessed against inclusion and exclusion criteria. Ten participants were excluded due to violations initiated by others than the caregiver-infant dyad or based on their low coded reunion time, so 157 participants were included in the study (Figure 1).

Demographic characteristics of the preterm (n= 82) and term (n = 75) groups are displayed in Table 1. Mean (SD) GA for the preterm group was 29.49 (2.14) weeks, mean birthweight was 1325 (389) g and mean days in NICU were 53.89 (27.93). There were no differences in sex or ethnicity between the preterm and the term group. The preterm group had a significantly lower SIMD rank compared to the term group (mean (SD) of 4219 (1950) and 4787 (1813), respectively; W = 3639, p-value < .05), which indicates higher deprivation in the preterm group.

**Table 1.**
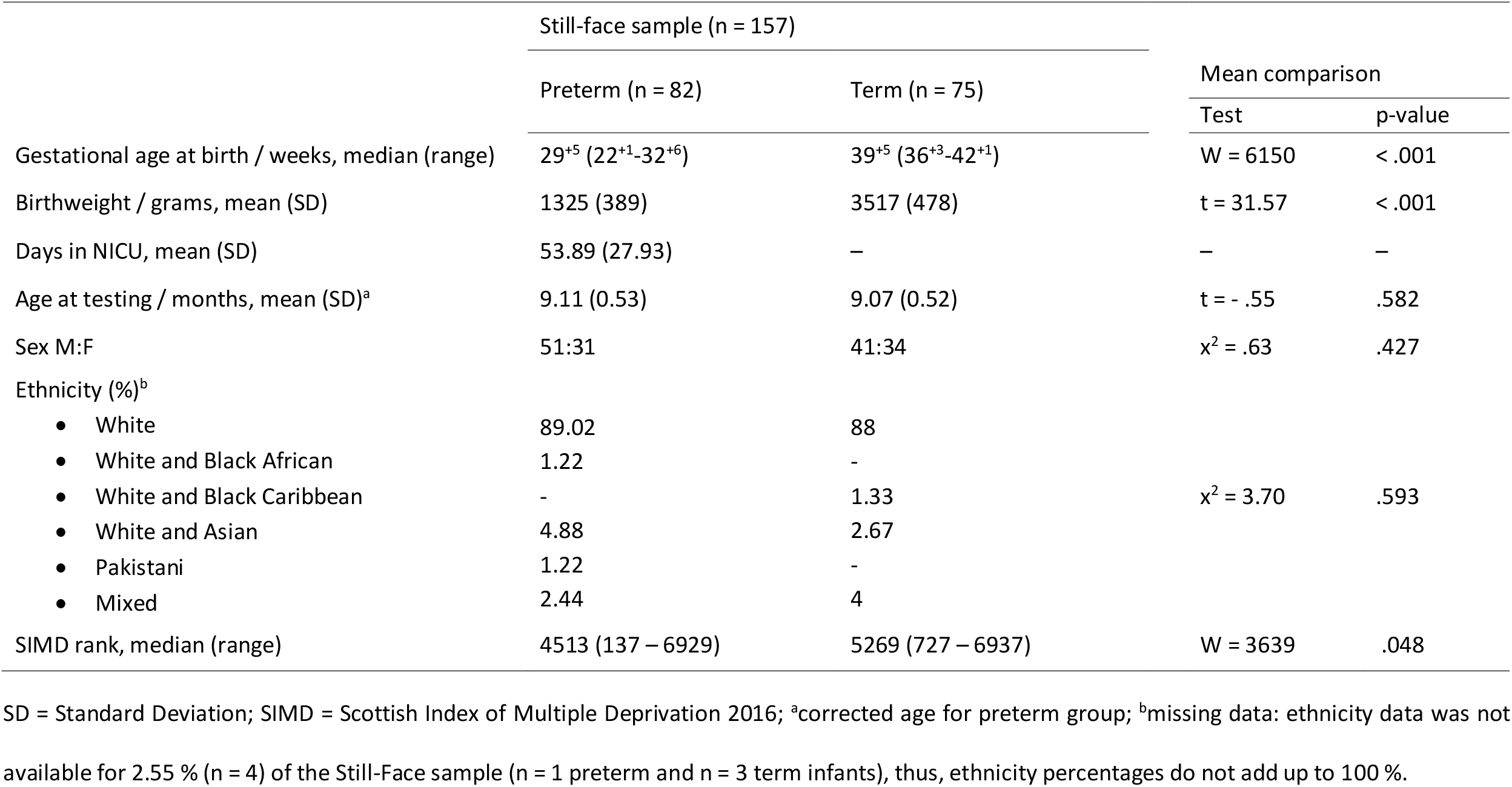
Demographic characteristics by group.

|  | Still-face sample (n = 157) |  | Mean comparison |  |
| --- | --- | --- | --- | --- |
|  | Preterm (n = 82) | Term (n = 75) | Test | p-value |
| Gestational age at birth / weeks, median (range) | 29<sup>+5</sup> (22<sup>+1</sup>-32<sup>+6</sup>) | 39<sup>+5</sup> (36<sup>+3</sup>-42<sup>+1</sup>) | W = 6150 | < .001 |
| Birthweight / grams, mean (SD) | 1325 (389) | 3517 (478) | t = 31.57 | < .001 |
| Days in NICU, mean (SD) | 53.89 (27.93) | — | — | — |
| Age at testing / months, mean (SD)<sup>a</sup> | 9.11 (0.53) | 9.07 (0.52) | t = - .55 | .582 |
| Sex M:F | 51:31 | 41:34 | $\chi^2 = .63$ | .427 |
| Ethnicity (%)<sup>b</sup> |  |  |  |  |
| • White | 89.02 | 88 | $\chi^2 = 3.70$ | .593 |
| • White and Black African | 1.22 | - |  |  |
| • White and Black Caribbean | - | 1.33 |  |  |
| • White and Asian | 4.88 | 2.67 |  |  |
| • Pakistani | 1.22 | - |  |  |
| • Mixed | 2.44 | 4 |  |  |
| SIMD rank, median (range) | 4513 (137 – 6929) | 5269 (727 – 6937) | W = 3639 | .048 |

SD = Standard Deviation; SIMD = Scottish Index of Multiple Deprivation 2016; <sup>a</sup>corrected age for preterm group; <sup>b</sup>missing data: ethnicity data was not available for 2.55 % (n = 4) of the Still-Face sample (n = 1 preterm and n = 3 term infants), thus, ethnicity percentages do not add up to 100 %.

### 6.2 Attachment behaviors in the Still-Face Paradigm in preterm vs term infants

Raw scores of the HD, NFF and AA scales per group are reported in Table 2. There were no significant differences in HD raw scores (W = 3271, p-value = .49, r = .06), or AA raw scores (W = 3503, p-value = .13, r = .12) of preterm compared to term infants. NFF raw scores were significantly lower in preterm compared to term infants (W = 2503, p-value = .02, r = .18).

**Table 2.** Attachment dimensions and categories in preterm and term infants.

|  |  |  | Still-face sample (n = 157) |  |  |  |  |
| --- | --- | --- | --- | --- | --- | --- | --- |
|  |  |  |  |  | Comparison |  |  |
|  |  |  | Preterm (n = 82) | Term (n = 75) | Test | p-value | Effect size estimate |
| Existing categorization | Attachment behaviors | HD raw score, median (range) | .54 (.03 – .88) | .56 (0 – .95) | W = 3271 | .492 | r = .06 |
|  |  | NFF raw score, median (range) | 1 (.49 – 1) | 1 (.23 – 1) | W = 2503 | .024 | r = .18 |
|  |  | AA raw score, median (range) | .28 (.01 – .77) | .31 (0 – .83) | W = 3503 | .133 | r = .12 |
| | | Secure, n (%) | 19 (23.17) | 22 (29.33) | $\chi^2 = 1.75$ | .782 | Cramér's V = .11 |
|  |  | Avoidant, n (%) | 39 (47.56) | 31 (41.33) |  |  |  |
|  |  | Resistant, n (%) | 1 (1.22) | 2 (2.67) |  |  |  |
|  |  | Disorganized, n (%) | 20 (24.39) | 16 (21.33) |  |  |  |
|  |  | Unscorable, n (%) | 3 (3.66) | 4 (5.34) |  |  |  |
| Principal component and clustering analyses | Attachment principal component scores | PC1, median (range) | .07 (-3.99 – 2.66) | .34 (-5.37 – 3.56) | W = 3220 | .612 | r = .04 |
|  |  | PC2, median (range) | -.24 (-1.65 – 2.31) | .13 (-1.68 – 2.50) | W = 3816 | .009 | r = .21 |
| | | I, n (%) | 7 (8.53) | 16 (21.33) | $\chi^2 = 6.90$ | .032 | Cramér's V = .21 |
|  |  | II, n (%) | 48 (58.54) | 31 (41.33) |  |  |  |
|  |  | III, n (%) | 27 (32.93) | 28 (37.33) |  |  |  |

AA = Attentive-Avoidant scale; HD = Happy-Distressed scale; NFF = Not fretful-Fretful scale; PC1 = first principal component; PC2 = second principal component; SD = standard deviation.

### 6.3 Attachment styles in preterm vs term infants

Distributions of attachment styles per group are reported in Table 2. There were no significant differences in the distribution of attachment styles between preterm and term infants (x^2^ = 1.75, p-value = .78, Cramér’s V = .11). Prematurity was not significantly associated with any attachment style (Table S1).

### 6.4 Attachment principal components in preterm vs term infants

To compare data-driven attachment dimensions between preterm and term infants, two PCs were retained which explained 86.95 % of the total variance. The main contributor of the first PC (PC1) were HD raw scores (i.e., “emotional” PC1) whereas the main contributor of the second PC (PC2) were AA raw scores (i.e., “attentional” PC2; Table S2). Attachment PC scores per group are reported in Table 2. There were no significant differences in emotional PC1 scores of preterm compared to term infants (PC1; W = 3320, p-value = .61, r = .04). Attentional PC2 scores were significantly lower in preterm compared to term infants (W = 3816, p-value < .01, r = .21).

### 6.5 Attachment clusters in preterm vs term infants

Hierarchical clustering suggested three clusters as the best fit to the data (Table S3). Infants in Cluster I showed higher distress (lower HD raw scores), especially in the second half of the reunion episode, higher fretfulness (lower NFF raw scores) and lower attentiveness to the caregiver (lower AA raw scores). Infants in Cluster II were neutral during the reunion episode (medium and similar HD raw scores across the episode), showed lower fretfulness (higher NFF raw scores) and lower attentiveness. Infants in Cluster III showed lower distress during the reunion episode (higher HD raw scores across the episode), lower fretfulness and higher attentiveness to the caregiver (higher AA raw scores). All clusters demonstrated high stability (average Rand Indices across iterations ranged from .88 to 1; Figure S3).

To explore the correspondence between attachment styles and clusters, the distribution of attachment styles within clusters were investigated (Table S4). There was a significant correspondence between attachment styles and clusters (x^2^ = 137.59, p-value < .001, Cramér’s V = .66, Figure 2), indicating good cluster validity in capturing attachment style. Cluster I captured infants assigned resistant, disorganized or unscorable attachment styles, Cluster II predominantly captured infants assigned an avoidant attachment style, and Cluster III predominantly captured infants assigned a secure attachment style.

**Figure 2.**
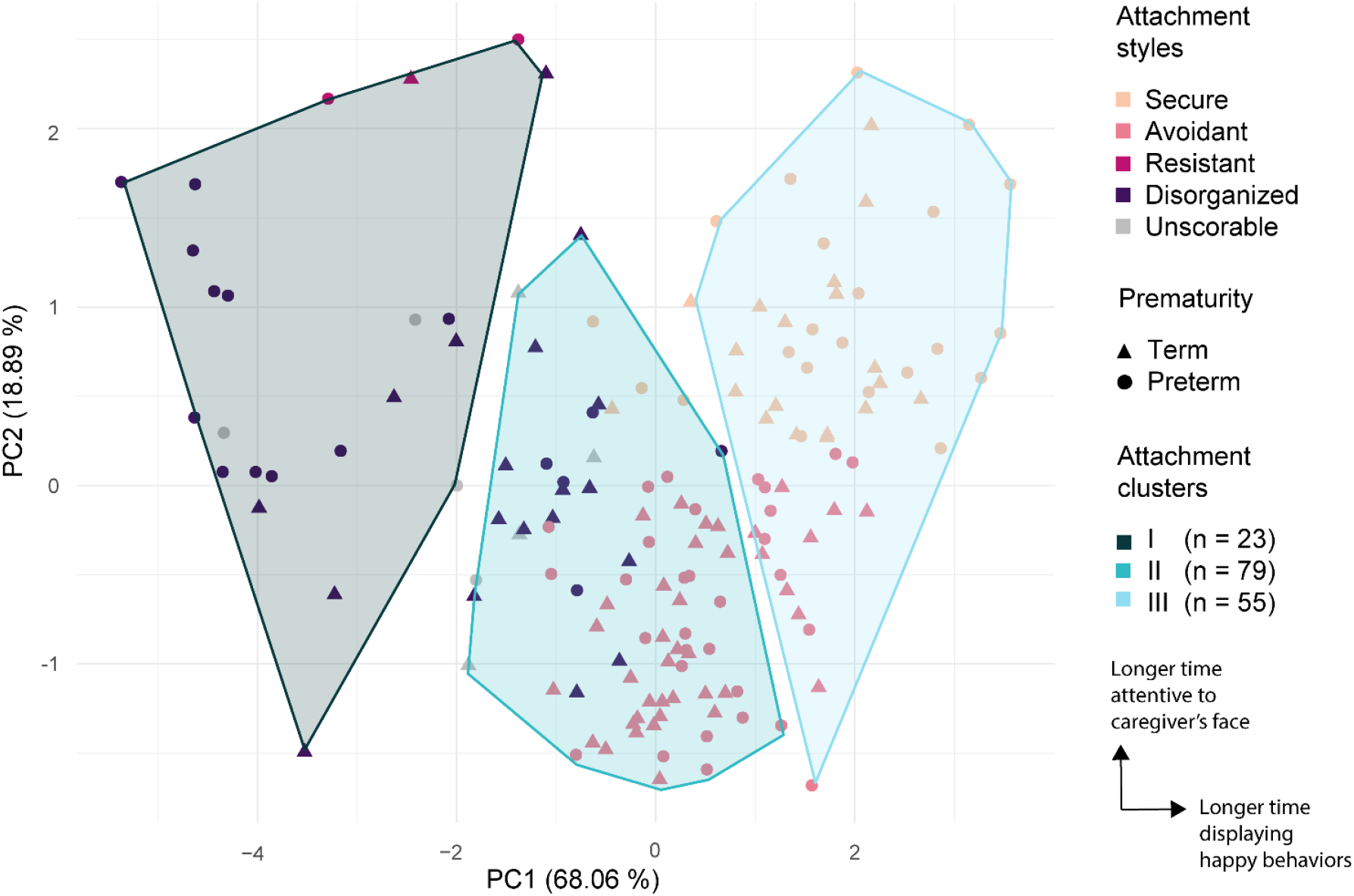
Factor map of attachment styles within attachment clusters. Contributions of the first (PC1, “emotional”) and second (PC2, “attentional”) principal components to the total variance are shown in brackets on the X and Y axes. Each point represents one infant (n = 157), color and shape indicate the assigned attachment style and prematurity of the infant, respectively. Shaded areas denote attachment clusters. See Table 2 and Table S3 for further details.

Distributions of attachment clusters per group are reported in Table 2. There were significant differences in the distribution of attachment clusters between preterm and term infants (x^2^ = 6.90, p-value = .03, Cramér’s V = .21). Prematurity was not significantly associated with any attachment cluster (Table S4).

### 6.6 Correlations between attachment dimensions and demographic characteristics at birth

Correlations between attachment dimensions and potentially confounding factors were explored. These factors included GA as a proxy measure for individual differences in prematurity, and SIMD rank since it significantly differed between preterm and term infants (Table 1).

GA did not significantly correlate with any attachment dimension. There was a positive correlation between GA and attentional PC2 scores that did not reach statistical significance (Spearman’s rho = .15, p-value= .06, Figure 3, Table S4); i.e. infants with lower GA may have lower attentional PC2 scores during the reunion episode of the SFP. Lower SIMD rank correlated with higher NFF raw scores (Spearman’s rho = -.18, p-value = .02; Figure 3, Table S5); i.e. infants from more deprived areas spent shorter time displaying fretful behaviors during the reunion episode of the SFP.

**Figure 3.**
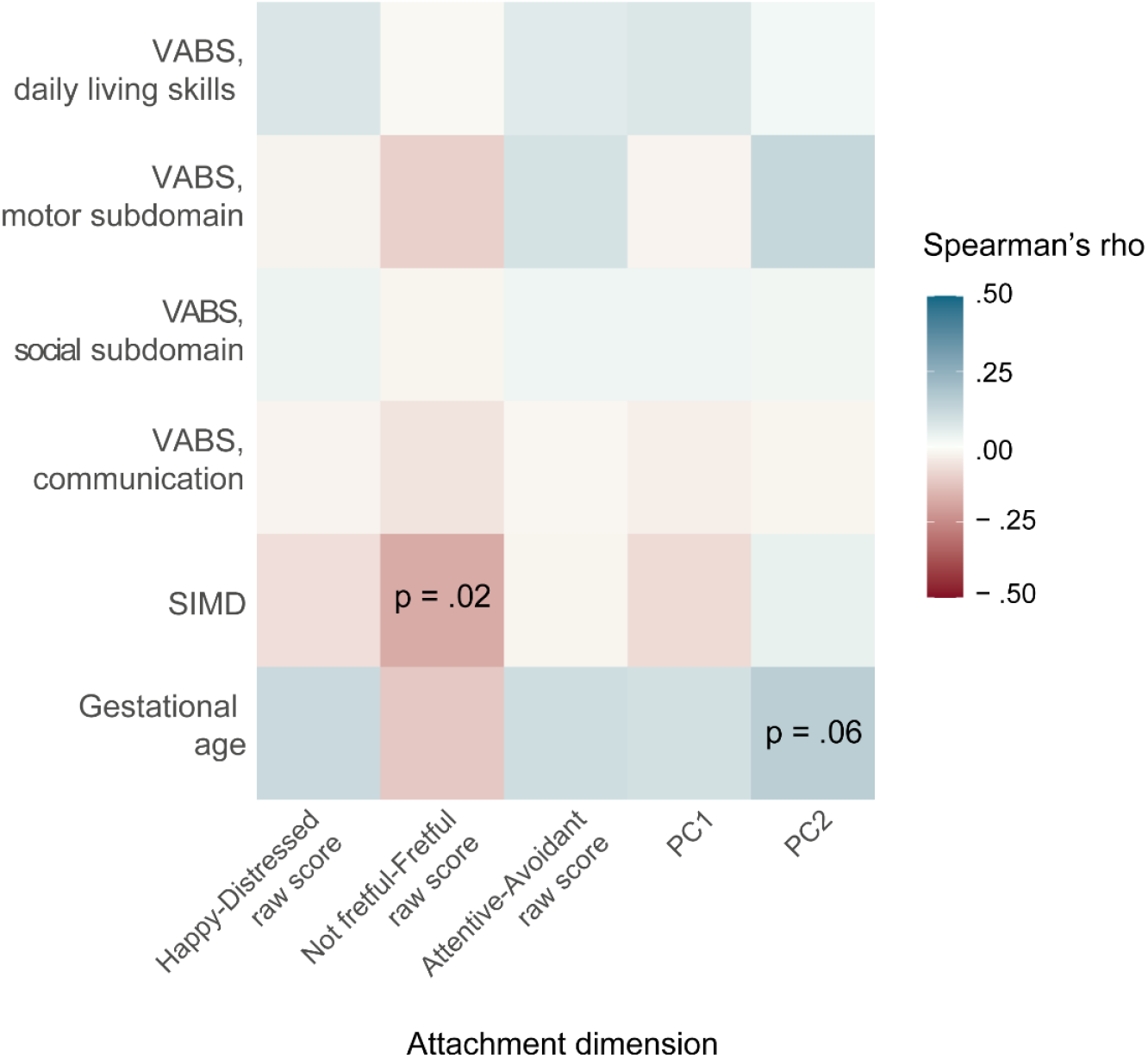
Heatmap showing correlations between attachment dimensions and demographic factors or behavioral development. Participants with missing data were excluded to perform a complete-case analysis (n = 157 for gestational age, SIMD rank, and VABS social, communication and motor subdomain; n = 156 for VABS daily living skills). Color indicates Spearman’s rho for each condition. P = p-value; PC1 = first principal component (i.e., “emotional” first principal component); PC2 = second principal component (i.e., “attentional” second principal component); VABS = Vineland Adaptive Behavior Scales (sum of raw scores per subdomain). See Table S4 for further details.

### 6.7 Cross-sectional associations between attachment dimensions and behavioral development at nine months of age

Since attachment behaviors could be confounded by infant behavioral development, correlations between attachment dimensions and infants’ adaptive behaviors were explored. There were no significant correlations between attachment dimensions and adaptive behaviors at nine months of age (Figure 3, Table S5).

### 6.8 Confounder analyses

NFF raw scores, attentional PC2 scores and the distribution of attachment clusters significantly differed between preterm and term infants. Prematurity did not associate with individual attachment clusters (Table S4), so no adjusted analysis was conducted to compare the distribution of attachment clusters between preterm and term infants. SIMD rank significantly correlated with infant NFF raw scores, and NFF raw scores were the second contributor to attentional PC2 scores (Table S2) so SIMD rank was controlled for in adjusted analyses comparing NFF raw scores and attentional PC2 scores between preterm and term infants. Controlling for SIMD rank did not change the direction of significant associations between prematurity and NFF raw scores or attentional PC2 scores (Table S6).

## 7 Discussion

This study found that some aspects of attachment significantly differed between preterm and term infants: preterm infants displayed lower fretfulness and PC2 scores during the reunion episode of the SFP, and might more frequently be assigned to attachment Cluster II. PC2 scores were based on coded attentiveness to the caregiver (with a minor contribution of coded fretfulness), so these were therefore designated “attentional”. Cluster II was characterized by more neutral, less fretful and less attentive responses during the reunion episode of the SFP. These results suggest preterm infants showed a more neutral and avoidant emotional response during the reunion episode in the observed samples. There were no significant differences in distress, attentiveness to the caregiver, PC1 scores (reflecting on coded distress, with a minor contribution of coded fretfulness) or in the distribution of traditional attachment styles between preterm and term infants. Gestational age at birth and behavioral development did not significantly correlate with any attachment dimension, and SIMD rank negatively correlated with infant fretfulness.

Reduced fretfulness may lead to reduced negative affect in preterm infants, as shown previously in a subset of this study sample (Ginnell et al., 2022). Preterm infants were found to display less negative affect compared to term infants during the still-face but not during the reunion episode of the SFP (Ginnell et al., 2022), so a larger sample in this study may have helped identify behavioral differences during the reunion episode. Differences in attentiveness to the caregiver and fretfulness (r = .12, r = .18; respectively) can explain significantly lower attentional PC2 scores in preterm compared to term infants during the reunion episode (see table S1 for PC2 loadings). A positive correlation between gestational age at birth and attentional PC2 scores in the whole sample (Figure 3, Table S4) indicates infants with lower gestational age at birth might display less attentiveness to the caregiver and less fretfulness during the SFP, suggesting a continuous relationship between infant gestational age at birth, attentiveness and fretfulness.

Significant differences in fretfulness did not lead to differences in attachment style distribution between preterm and term infants using an existing SFP categorization system (Williams and Turner, 2020). These results agree with other studies reporting that preterm infants are comparable in their attachment classifications to term infants from 12 months of age (Rode et al., 1981, Frodi and Thompson, 1985, Goldberg et al., 1986, Plunkett et al., 1986, Easterbrooks, 1989, Butcher et al., 1993, Brisch et al., 2005). By contrast, there were significant differences in attachment cluster distribution between preterm and term infants using a sample-specific data-driven approach. Although more preterm infants were assigned to Cluster II, prematurity did not significantly associate with Cluster II (Table S4) perhaps due to insufficient statistical power to detect a small effect size by logistic regression analyses. Since Cluster II predominantly captured infants assigned an avoidant attachment style, our results are consistent with the reported higher frequencies of avoidant attachment in preterm infants compared to term infants from 12 months of age (Fuertes et al., 2022). Our findings partially explain inconsistent results in studies investigating whether infant attachment is disrupted by preterm birth: behavioral differences may not always be captured by traditional approaches for categorizing attachment. Data-driven approaches do not outperform traditional categorization systems, but can help identify and understand subtle behavioral differences in attachment in preterm infants from 9 months of age.

Our results do not support the reported higher frequencies of disorganized attachment style (Wolke et al., 2014), and disorganized attachment scores (Zengin Akkus et al., 2021) in preterm infants compared to term infants from 12 months of age. Additionally, more insecure resistant attachment (López-Maestro et al., 2017, Sierra-García et al., 2018) and anxious attachment scores (Zengin Akkus et al., 2021) have been found in preterm infants at 18 months of age, whereas only three infants were assigned a resistant attachment style in this study. Interestingly, most studies using the Strange Situation Procedure show that approximately two thirds of preterm infants are assigned a secure attachment style from 12 months of age (Korja et al., 2012). We found a low prevalence of secure attachment style in this study using the SFP (Table 2), in line with Williams and Turner who reported similar findings in a cohort of term infants at seven months of age (2020). Infants need to be highly attentive to the caregiver to be classified as secure, so low infant attentiveness (i.e., low AA raw scores) may account for the low prevalence of secure attachment in this study.

In this sample, one demographic factor associated with infants’ attachment. Socioeconomic deprivation negatively correlated with infants’ fretfulness during the reunion episode of the SFP, suggesting infants strategies to re-engage with caregivers after separation may vary across socioeconomic backgrounds. Although socioeconomic deprivation did not confound significant associations between prematurity and attachment in this sample, prematurity could be more likely to impact attachment when it is associated with family socioeconomic disadvantage (Fuertes et al., 2022, Wille, 1991). Future studies could explore interactions between prematurity, socioeconomic deprivation and attachment further. Interestingly, no aspect of behavioral development correlated with attachment, suggesting the development of attachment relationships may be independent of the development of adaptive behaviors in infancy.

### 7.1 Limitations, strengths and future directions

This study relied on coding infant behavior from video and did not include maternal measures, for instance maternal sensitivity, which has been shown to predict attachment organization in term infants (Wolke et al., 2014). However, this work was strengthened by use of a data-driven approach to avoid reliance on expected attention capacities in a cohort of preterm infants, who may present with different levels of attention maturation (Ginnell et al., 2021). Despite the inherent challenges of coding infant behavior (e.g., tracking the infant’s gaze is difficult in distressed infants who cry and close their eyes for some time), we reached good reliability between coders. We included gestational age at birth to account for individual differences in prematurity and explored links between attachment and possible confounders, including demographic factors that differed between the preterm and term group and behavioral development. Caregiving practices and infant social networks can differ substantially across cultures (Schmidt et al., 2021), so future studies could explore whether these findings are generalizable to other populations.

### 7.2 Conclusion

Data reveal subtle attachment differences between preterm and term infants at nine months of age, that may not always be captured by traditional approaches for categorizing attachment. Findings suggests that caregiver-infant attachment relationships may not be fully resilient to the effects of prematurity on the developing infant, but this depends on how attachment is measured. Our results highlight putative links between socioeconomic deprivation and infant attachment that warrant further study.

## 8. Key points and relevance

- The adverse impact of preterm birth on socioemotional outcomes might be mitigated by secure infant attachment, but this could be affected by early exposure to extrauterine life.
- We used an existing categorization system and a data-driven approach on infant behaviors coded from the Still-Face Paradigm to test whether preterm birth leads to differences in infant attachment, and identified variables that associated with infant attachment.
- Preterm infants exhibited a more neutral and avoidant response that was not fully captured by an existing system to categorize attachment.
- Attachment relationships may not be fully resilient to the effects of prematurity on the developing infant, but this depends on how attachment is measured.
- Socioeconomic deprivation negatively correlated with infants’ fretfulness, which warrants further study.

## Data Availability

Data availability is governed in accordance with the Theirworld Edinburgh Birth Cohort data access and collaboration policy, which is available on request

## 11 Acknowledgements

The authors are grateful to the families who participated in this research.

## Abbreviations

AA: Attentive-Avoidant scale
GA: Gestational age at birth
HD: Happy-Distressed scale
ICC: Intraclass correlation coefficient
NFF: Not fretful-Fretful scale
NICU: Neonatal Intensive Care Unit
PC: Principal component
PC1: First principal component
PC2: Second principal component
SD: Standard deviation
SIMD: Scottish Index of Multiple Deprivation
SFP: Still-Face Paradigm
VABS: Vineland Adaptive Behavior Scales

