## Supporting information for "Early life attachment in term and preterm infants"

**Appendix S1.** Exclusion criteria.

**Appendix S2.** R packages used for statistical analyses.

**Appendix S3.** Cluster robustness.

**Figure S1.** Comparison of Vineland's Adaptive Behavior Scales scores.

**Figure S2.** Reproducibility of bootstrapping based on iteration number.

**Table S1.** Relationship between prematurity and individual attachment styles.

**Table S2.** Input variable loadings to principal components.

**Table S3.** Input variables within attachment clusters.

**Table S4.** Distribution of attachment styles within attachment clusters.

**Table S5.** Relationship between prematurity and individual attachment clusters.

**Table S6.** Correlations between attachment dimensions and demographic factors or behavioral development.

**Table S7.** Relationship between prematurity and attachment adjusting for confounders.

**References.**

**Appendix S1. Exclusion criteria.**

We identified three main factors that could change the predictive behavioral responses of the infants during the Still Face Paradigm (SFP) or affect the video coding process:

- SFP procedural violations: During the SFP, the possibility existed that the caregiver or researchers violated the procedure. Since only the first reunion episode was coded, only violations that happened before the first reunion episode were considered relevant for this study. During the first play or reunion episodes, violations included the use of objects not inherent to the face-to-face setting (pacifier, bottle or toys). During the still-face episode, violations included the caregiver turning away, touching or talking to the baby. Other relevant violations in this study included the infant's early termination of the reunion episode (due to distress), interruptions by researchers or family members, researchers not concealed from view of the infant and caregiver, or infants being obscured by the camera.
- Wear of sensors: a subset sample of infants wore movement sensors on their wrists, ankles and torso as part of a sub-group analysis within the cohort protocol.
- Available coded reunion time: coded reunion time refers to the duration of the codable time during the reunion episode, in other words, the total number of seconds of the reunion time minus the number seconds of "unscorable" behavior (i.e. when the infant's face was not visible; Williams and Turner, 2020). The available coded reunion time should be long enough to represent the infant's response to the SFP.

Cases where infants could not terminate the reunion episode due to distress (early termination) or where the caregiver-infant dyad initiated the violation were considered to capture the clinical heterogeneity of the sample. Therefore, these participants were not excluded. However, cases where others than the caregiver-infant dyad i.e. researchers and other family members initiated the violation (researchers not concealed from view of the infant and caregiver, siblings present in room, etc.) were excluded. There were no differences in the frequency of infants wearing sensors between the preterm and the term group (n = 20 of all participants providing codable videos wore sensors; $x^2$ = .03, p = .87), so no participants were excluded for this reason. Finally, the shortest duration of SFP episodes at nine months of age was 30 seconds in previous studies (Striano et al., 2005; see Mesman et al., 2009 for a review on the duration of the SFP). Thus, cases where the coded reunion time was lower than 30 seconds were also excluded.

**Appendix S2. R packages used for statistical analyses.**

| Analysis | R package name | Citation |
|---|---|---|
| Principal component analysis | ‘factoextra’ | (Kassambara and Mundt, 2019) |
| Agglomerative hierarchical clustering | ‘FactoMineR’ | (Lê et al., 2008) |
| Effect size estimation: Cramér’s V, Cohen’s d, | ‘lsr’ | (Navarro and Navarro, 2021) |
| Effect size estimation: r (rank biserial correlation) | ‘rstatix’ | (Kassambara, 2021) |
| Inter-reliability scores: Intraclass correlation coefficient (Sparrow et al.), percentage of agreement and Cohen’s kappa. | ‘irr’ | (Gamer, 2010) |
| Cluster robustness: bootstrapping. | ‘bluster’ | (Lun and Hicks, 2022) |
| Multinomial logistic regression | ‘nnet’ | (Ripley et al., 2016) |

**Appendix S3. Cluster robustness.**

Data-driven approaches can sometimes provide cluster solutions despite weak cluster separation, so it is important to determine whether a cluster solution is stable and should be retained. Following cluster analysis, bootstrapping was used to assess the clusters stability for several reasons: 1) it has recently been used to assess cluster stability of clinical phenotypes in preterm infants (Matsushita et al., 2022), 2) we aimed to draw all of the new data in replicated datasets from the same sample (King, 1997), and 3) we aimed to keep the sample size and statistical power as constant as possible across replicated datasets. In-silico replicated datasets were generated from the original dataset and hierarchical clustering was performed in all replicates (B = 500 iterations). Since there is no consensus to determine how many iterations are required for a stable bootstrap estimate (see an example in Rosenfeld et al., 2017), the number of iterations was decided based on reproducibility of the Rand index values per pair of reference and alternative clusters across iterations over n = 3 trials. The distribution of values became stable after 500 iterations (Figure S2). Alternative clusters were compared to original clusters by computing the Rand index values for each pair of clusters on each iteration, and the matrix of the average Rand index values across iterations for each pair of clusters was reported. A matrix is reported as it is more interpretable than a single value (such as the adjusted Rand Index), and it focuses on the relevant differences between clusters, helping to determine which aspects of clustering are more stable. For example, Cluster I and II may be well separated but Cluster I and III may not be, which is not accurately represented by a single stability measure for Cluster I.

**Figure S1. Comparison of Vineland's Adaptive Behavior Scales scores.**

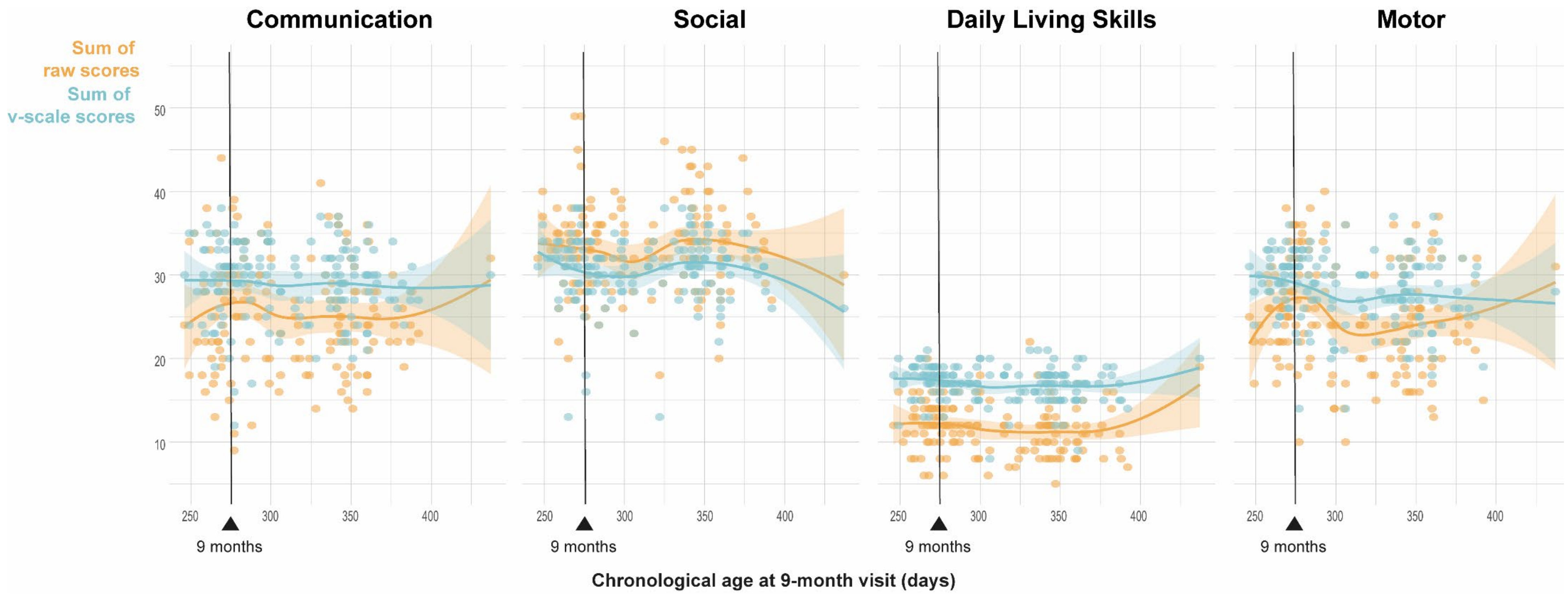


*Parents were interviewed at the 9-month visit to complete the Vineland's Adaptive Behavior Scales (VABS) questionnaire. Plots show data from n = 156 participants, with available data for all subdomains. The X axis represents infant chronological age at the 9-month visit (days), the Y axis indicates the sum of raw scores and v-scale scores per behavioral subdomain. Raw scores represent the total number of items (adaptive behaviors) infants present at the time of assessment, while v-scale scores are calculated by scaling raw scores (mean = 15, standard deviation = 3) according to the infant's corrected age at the time of assessment.*

**Figure S2. Reproducibility of bootstrapping based on iteration number.**

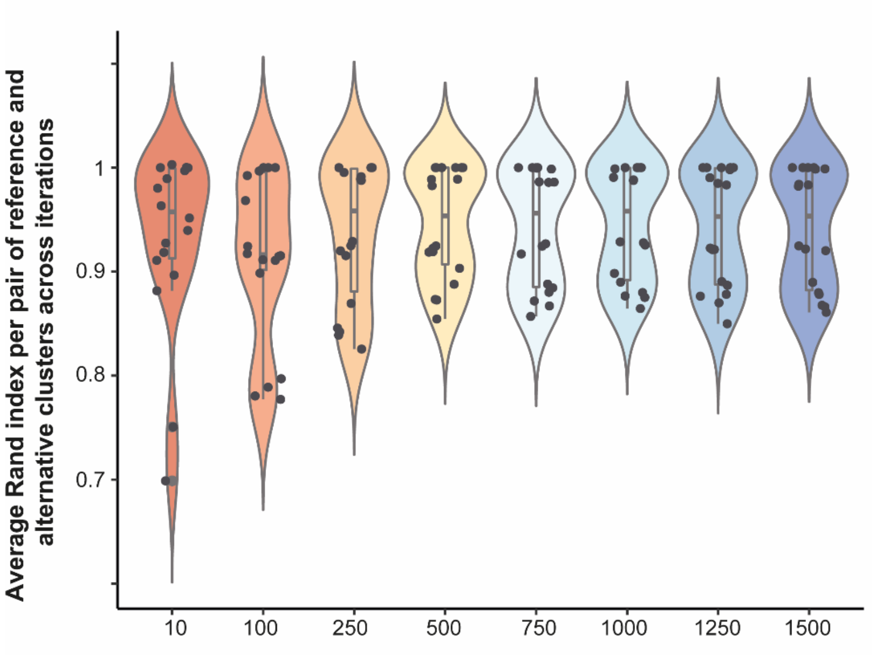


*Violin plots showing the distribution of average Rand index values per pair of reference and alternative clusters across different numbers of iterations. Each test (number of iterations) was repeated three times (n = 3 trials) to evaluate the reproducibility of results: the distribution of values became stable after 500 iterations.*

**Figure S3. Cluster stability.**

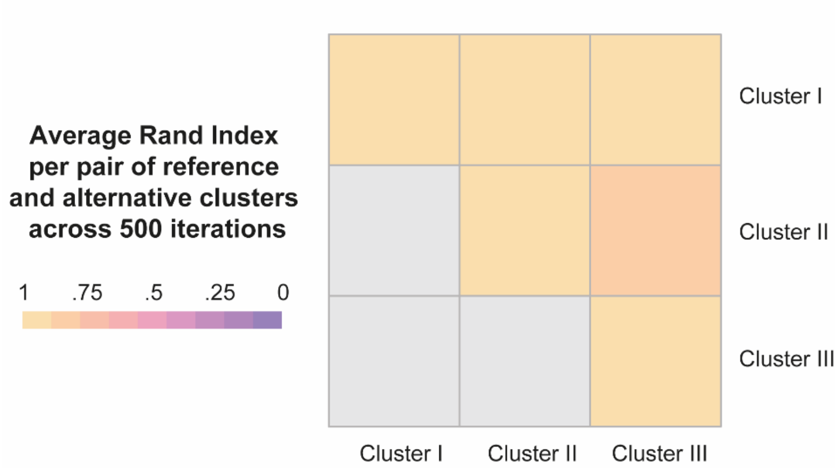


*High on-diagonal values indicate that the corresponding cluster remains coherent in the bootstrap replicates, while high off-diagonal values indicate that the corresponding pair of clusters are still separated in the replicates.*

**Table S1. Relationship between prematurity and individual attachment styles.**

| Attachment styles | Estimate | Standard error | RRR | z | p-value |
|---|---|---|---|---|---|
| Avoidant | | | | | |
| (Intercept) | .34 | .28 | 1.41 | 1.23 | .219 |
| Prematurity | .38 | .39 | 1.46 | .95 | .341 |
| Resistant | | | | | |
| (Intercept) | -2.40 | .74 | .09 | -3.25 | **.001** |
| Prematurity | - .55 | 1.26 | .58 | - .43 | .665 |
| Disorganised | | | | | |
| (Intercept) | - .32 | .33 | .73 | - .97 | .332 |
| Prematurity | .37 | .46 | 1.45 | .81 | .420 |
| Unscorable | | | | | |
| (Intercept) | -1.70 | .54 | .18 | -3.14 | **.002** |
| Prematurity | - .14 | .77 | .87 | - .17 | .864 |

*Multivariate logistic regression was used with secure attachment and term-born as reference groups. RRR = relative risk ratio. P-values < .05 are shown in bold.*

**Table S2. Input variable loadings to principal components.**

| | PC1 | PC2 | PC3 | PC4 | PC5 |
|---|---|---|---|---|---|
| Total HD raw score | 28.49 | .04 | 1.88 | 6.63 | 62.96 |
| First-half HD raw score | 23.40 | .03 | 57.69 | 1.22 | 17.66 |
| Second half HD raw score | 23.98 | .36 | 21.97 | 34.32 | 19.37 |
| Total NFF raw score | 18.86 | 16.69 | 14.03 | 50.42 | .01 |
| Total AA raw score | 5.28 | 82.88 | 4.44 | 7.40 | .00 |

*Loadings are provided as percentages. AA = Attentive-Avoidant scale; HD = Happy-Distressed scale; NFF = Not fretful-Fretful scale.*

**Table S3. Input variables within attachment clusters.**

| | Attachment clusters | | |
|---|---|---|---|
| | I<br>(n = 23) | II<br>(n = 79) | III<br>(n = 55) |
| Total HD raw score, median (range) | .14 (0 – .40) | .58 (.26 – .71) | .72 (.52 – .95) |
| First-half HD raw score, median (range) | .19 (0 – .63) | .52 (.10 – .73) | .75 (.50 – .96) |
| Second half HD raw score, median (range) | .05 (0 – .39) | .51 (.03 – .77) | .68 (.45 – 1) |
| Total NFF raw score, median (range) | .56 (.23 – 1) | 1 (.72 – 1.00) | 1 (.88 – 1) |
| Total AA raw score, median (range) | .22 (.01 – .65) | .18 (.00 – .49) | .47 (.01 – .83) |

*AA = Attentive-Avoidant scale; HD = Happy-Distressed scale; NFF = Not fretful-Fretful scale.*

**Table S4. Distribution of attachment styles within attachment clusters.**

| | | Attachment clusters | | | Comparison | | |
|---|---|---|---|---|---|---|---|
| | | I (n = 23) | II (n = 79) | III (n = 55) | Test | p-value | Effect size estimate |
| Attachment styles | Secure, n (%) | 0 (0) | 4 (5.06) | 37 (67.27) | $x^2$ = 137.59 | < .001 | Cramér's V = 0.66 |
| | Avoidant, n (%) | 0 (0) | 52 (65.83) | 18 (32.73) | | | |
| | Resistant, n (%) | 3 (13.04) | 0 (0) | 0 (0) | | | |
| | Disorganized, n (%) | 17 (73.91) | 19 (24.05) | 0 (0) | | | |
| | Unscorable, n (%) | 3 (13.04) | 4 (5.06) | 0 (0) | | | |

**Table S4. Relationship between prematurity and individual attachment clusters.**

| Attachment clusters | Estimate | Standard error | RRR | z | p-value |
|---|---|---|---|---|---|
| I | | | | | |
| (Intercept) | - .56 | .31 | .57 | -1.79 | .074 |
| Prematurity | - .79 | .53 | .45 | -1.50 | .134 |
| II | | | | | |
| (Intercept) | .10 | .26 | 1.11 | .39 | .696 |
| Prematurity | .47 | .35 | 1.61 | 1.34 | .182 |

*Multivariate logistic regression was used with cluster III and term-born as reference groups. RRR = relative risk ratio. P-values < .05 are shown in bold.*

**Table S5. Correlations between attachment dimensions and demographic factors or behavioral development.**

| Variables | | Spearman's rho | p-value |
|---|---|---|---|
| Gestational age at birth | Total HD raw score | .12 | .149 |
| | Total NFF raw score | - .12 | .149 |
| | Total AA raw score | .10 | .191 |
| | PC1 | .09 | .240 |
| | PC2 | .15 | .056 |
| SIMD rank | Total HD raw score | - .07 | .405 |
| | Total NFF raw score | - .18 | **.025** |
| | Total AA raw score | - .02 | .792 |
| | PC1 | - .07 | .359 |
| | PC2 | .04 | .598 |
| VABS social subdomain[a] | Total HD raw score | .04 | .663 |
| | Total NFF raw score | - .02 | .785 |
| | Total AA raw score | .03 | .723 |
| | PC1 | .03 | .705 |
| | PC2 | .02 | .762 |
| VABS communication subdomain[a] | Total HD raw score | - .02 | .762 |
| | Total NFF raw score | - .06 | .489 |
| | Total AA raw score | - .02 | .827 |
| | PC1 | - .03 | .682 |
| | PC2 | - .02 | .784 |
| VABS motor subdomain[a] | Total HD raw score | - .03 | .754 |
| | Total NFF raw score | - .10 | .224 |
| | Total AA raw score | .09 | .273 |
| | PC1 | - .02 | .766 |
| | PC2 | .12 | .123 |
| VABS daily living skills[a] | Total HD raw score | .08 | .325 |
| | Total NFF raw score | - .01 | .854 |
| | Total AA raw score | .06 | .437 |
| | PC1 | .07 | .358 |
| | PC2 | .02 | .781 |

*[a] Participants with missing data were excluded to perform a complete-case analysis (n = 157 for gestational age, SIMD, VABS social, communication and motor subdomain data, n = 156 for VABS daily living skills). AA = Attentive-Avoidant scale; HD = Happy-Distressed scale; NFF = Not fretful-Fretful scale; PC1 = first principal component; PC2 = second principal component; SIMD = Scottish Index of Multiple Deprivation 2016; VABS = Vineland Adaptive Behavior Scales (sum of raw scores per subdomain). P-values < .05 are shown in bold.*

**Table S6. Relationship between prematurity and attachment adjusting for confounders.**

| Attachment dimensions | Estimate | Standard error | t-value | p-value |
|---|---|---|---|---|
| NFF raw scores | | | | |
| (Intercept) | .92 | .04 | 23.23 | **< .001** |
| Prematurity | .07 | .03 | 2.39 | **.018** |
| SIMD rank | < .001 | < .001 | -1.30 | .195 |
| PC2 | | | | |
| (Intercept) | .23 | .23 | 1.03 | .304 |
| Prematurity | - .40 | .16 | -2.55 | **.012** |
| SIMD rank | < .001 | < .001 | - .14 | .890 |

*To compare attachment dimensions, generalized linear regression was used. NFF = Not fretful-Fretful scale; PC2 = second principal component. P-values < .05 are shown in bold.*
